# Is there an association between daytime napping, cognitive function and brain volume? A Mendelian randomisation study in the UK Biobank

**DOI:** 10.1101/2021.09.28.21264215

**Authors:** Valentina Paz, Hassan S. Dashti, Victoria Garfield

**Affiliations:** MRC Unit for Lifelong Health & Ageing, Institute of Cardiovascular Science, University College London, London, United Kingdom; Instituto de Psicología Clínica, Facultad de Psicología, Universidad de la República, Montevideo, Uruguay; Center for Genomic Medicine, Massachusetts General Hospital and Harvard Medical School, Boston, Massachusetts, United States of America; Broad Institute, Cambridge, Massachusetts, United States of America; Department of Anesthesia, Critical Care and Pain Medicine, Massachusetts General Hospital and Harvard Medical School, Boston, Massachusetts, United States of America

**Keywords:** Mendelian randomisation, Daytime napping, Cognitive function, Brain volume

## Abstract

Daytime napping has been associated with cognitive function and brain health in observational studies. However, it remains elusive whether these associations are causal. Using Mendelian randomisation (MR), we studied the relationship between habitual daytime napping and cognitive/structural brain outcomes. Data were from UK Biobank (UKB) (maximum *n*= 378 932; mean age= 57 years). Our exposure (daytime napping) was instrumented using 92 previously identified genome-wide, independent genetic variants (single-nucleotide polymorphisms, SNPs). Our cognitive outcomes were reaction time and visual memory; our neuroimaging outcomes were total brain volume and hippocampal volume (cm3). Inverse-variance weighted (IVW) MR was implemented, with sensitivity analyses including MR-Egger and the Weighted Median Estimator for horizontal pleiotropy. We also tested different daytime napping instruments (47 SNPs, 86 SNPs and 17 SNPs) to ensure the robustness of our results. Our main MR analysis (IVW) showed an association between genetic liability to habitual daytime napping and larger total brain volume (unstandardised ß=15.80 cm3, 95% confidence interval (CI)=0.25; 31.34), but not hippocampal volume (ß=-0.03 cm3, 95%CI=-0.13; 0.06). No associations were found between daytime napping and reaction time (expß=1.01, 95%CI=1.00; 1.03), or visual memory (expß=0.99, 95%CI=0.94; 1.05). Additional analyses with 47 SNPs (adjusted for excessive daytime sleepiness), 86 SNPs (excluding sleep apnoea) and 17 SNPs (no sample overlap with UKB) were largely consistent with our main findings. MR-Egger and Weighted Median Estimator approaches showed no evidence of horizontal pleiotropy. Overall, we observed evidence of an association between genetically-instrumented daytime napping and larger total brain volume, but no evidence of an association between habitual daytime napping and hippocampal volume, reaction time or visual memory. Future studies could focus on the associations between napping and other cognitive/brain outcomes, as well as replication of these findings using other datasets and methods.

**Key Messages:**

- Daytime napping has been linked with cognitive function and brain health in observational studies, but whether these links are causal remains elusive.
- Using Mendelian randomisation, we investigated the potential causal role of habitual daytime napping on cognitive and neuroimaging outcomes.
- We observed evidence of a modest causal association between habitual daytime napping and larger total brain volume, but not enough evidence to support associations with hippocampal volume, reaction time or visual memory.

## Introduction

Daytime napping, defined as brief daytime bouts of sleep [1], is a universal [2, 3] and prevalent behaviour [4], reported in approximately 30% of the British population [5]. Napping has been associated with multiple health outcomes [4, 6], including cognitive [7, 8] and structural brain outcomes [9]. Napping seems beneficial to performance on certain cognitive tasks [3, 10, 11]. These benefits arise immediately following a brief nap (e.g. five to 15 minutes) and can last between one to three hours. After a long nap (>30 min), a temporary deterioration of performance emerges, followed by improvements that can last up to a day [11]. However, some authors argue that individuals who frequently have a nap and those who never nap may differ in the benefits derived from napping, with the latter experiencing no benefits from it [3]. While recently more attention has been paid to napping, it remains elusive whether habitual daytime napping could be beneficial or detrimental for cognition [12]. In addition, the association between napping and brain volume is not well characterised, even though changes in brain volumes are strong candidates to explain variations in cognition [13, 14]. Moreover, as most studies about the relationship between napping and cognitive/brain health are observational, causal associations between both could not be drawn.

To overcome this limitation, Mendelian randomisation (MR) can be used, which is based on the analysis of genetic markers, to examine the possible causal associations between exposures and outcomes [15, 16]. Previous MR studies investigated the causal relationship between sleep and cognitive and structural brain outcomes. These studies reported that both short and long sleep duration are associated with poorer cognitive outcomes [17], long sleep duration is associated with increased cortical thickness [18], and different sleep traits are associated with a greater risk of neurodegenerative diseases [19–21]. Regarding napping, Anderson et al. (2021) found suggestive evidence that self-reported habitual daytime napping is associated with lower Alzheimer’s disease (AD) risk. However, no previous MR studies have investigated the association between daytime napping, cognitive outcomes and brain volumes. Given that the most pronounced decline during ageing occurs in reaction time and memory [23], and the high prevalence of cognitive impairment in the ageing population [24], the identification of modifiable risk factors is essential. Thus, the present study aimed to use MR to examine whether the relationship between genetic liability to daytime napping and: cognitive function and brain volumes might be causal.

## Methods

### Sample

The UK Biobank (UKB) cohort has been described in detail elsewhere [25]. Briefly, UKB recruited 500 000 males and females from the general UK population, aged 40-69 years at baseline (2006-2010). Although UKB recruited participants of distinct ancestries, those included in this study were of white European ancestry and retained if they had relevant (quality-controlled) genotype and phenotype data (*n*=378 932).

### Study design

Our exposure (SNPs_x_) sample overlapped with our cognitive function outcome sample (SNPs_y_) by 77%, but this was <10% for the neuroimaging outcomes. This is because the discovery genome-wide association study (GWAS) for the exposure under study was performed in UKB participants, which was also our analytical sample. However, below we detail in Sensitivity Analyses the strategy we undertook to mitigate this sample overlap.

### Genotyping and quality control (QC) in UKB

487 409 UKB participants were genotyped using one of two customised genome-wide arrays that were imputed to a combination of the UK10K, 1000 Genomes Phase three and the Haplotype Reference Consortium (HRC) reference panels, which resulted in 93 095 623 autosomal variants [26]. We then applied additional variant level QC and excluded genetic variants with: Fisher’s exact test <0.3, minor allele frequency (MAF) <1% and a missing call rate of ≥5%. Individual-level QC meant that we excluded participants with: excessive or minimal heterozygosity, more than 10 putative third-degree relatives as per the kinship matrix, no consent to extract DNA, sex mismatches between self-reported and genetic sex, missing QC information and non-European ancestry (based on how individuals had self-reported their ancestry and the similarity with their genetic ancestry, as per a principal component analysis of their genotype).

### Outcomes

#### Cognitive function measures

At baseline UKB administered a total of five cognitive assessments to all participants, via a computerised touch-screen interface, all of which are described in detail elsewhere [27]. For the purposes of this study and to maximise statistical power, we pragmatically chose *visual memory* and *reaction time*. For the *visual memory* task respondents were asked to correctly identify matches from six pairs of cards after they had memorised their positions. The number of incorrect matches (number of attempts made to correctly identify the pairs) was then recorded, with a greater number reflecting poorer visual memory. Reaction time (in milliseconds) was recorded as the mean time taken by participants to correctly identify matches in a 12-round game of the card game ‘Snap’. A higher score on this test indicated a slower (poorer) reaction time. Both of these variables were positively skewed and therefore, reaction time scores were transformed using the natural logarithmic function [ln(x)], whilst visual memory was transformed using [ln(x+1)].

#### Neuroimaging parameters

Structural brain magnetic resonance imaging (MRI) scans have been performed in a subsample of the UKB, using standard protocols (REF) **(see Supplementary Material)**. Here we had complete neuroimaging and genotype data for n=35,080 individuals. We analysed hippocampal volume (average of left + right hippocampal volume, cm3) and total brain volume (normalised for head size, cm3).

### Selection of genetic instruments

#### Main daytime napping genetic instrument

Daytime napping was instrumented using 123 genome-wide significant (*P*<5*10-8) genetic variants discovered in a recent genome-wide association study (GWAS) [28]. These variants were discovered in 452 633 UKB participants, based on the question ‘Do you have a nap during the day?’ administered at baseline, with possible responses Never/rarely, Sometimes and Usually (Prefer not to answer was coded as missing in the GWAS). Thirty-eight percent of UKB respondents reported that they ‘sometimes’ napped and 5% reported that they ‘usually’ have a nap. The 123 variants explain 1% of the variance in daytime napping. However, here we selected 92 of the 123 daytime napping SNPs, as we used linkage disequilibrium (LD) clumping in PLINK with r^2^<0.01 within 250kb. We then calculated the F-statistic which yielded F=41 using the Cragg-Donald formula [29]:

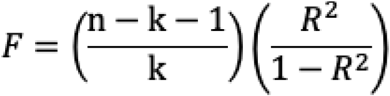

We harmonised the genetic variants between the exposure GWAS and our outcome sample by aligning effect alleles and we also excluded palindromic SNPs. Our instrument selection process is detailed in **Supplementary Figure S1**.

#### Additional daytime napping genetic instruments

We additionally partitioned the daytime napping instrument into two further sub-instruments: i) an 86-SNP instrument which consists of those SNPs that remained genome-wide significant when in the published GWAS the authors excluded individuals who had sleep apnoea (*n*=5553), ii) a 47-SNP instrument which comprised SNPs that remained genome-wide significant on adjustment for excessive daytime sleepiness. Using the formula F=(**β**^2^/SE^2^) to approximate average instrument strength for these additional instruments in sensitivity analyses, we calculated the F-statistic for each of these additional instruments, which yielded F=98.1 and F=47.0, respectively indicating good instrument strength.

### Statistical analyses

#### i. Main analyses

Using PLINK 2.0 we performed linear regressions between each of the daytime napping genetic variants and our outcomes, adjusting for 10 principal components to minimise issues of residual confounding by population stratification. For our MR analyses, inverse-variance weighted (IVW) MR was implemented, with standard sensitivity analyses including MR-Egger and the Weighted Median Estimator (WME). The IVW, also known as ‘conventional MR’ estimates the effect of an exposure (e.g. daytime napping) on a given outcome (e.g. visual memory/reaction time) by taking an average of the genetic variants’ ratio of variant-outcome (*SNP→Y*) to variant-exposure (*SNP→X*) association, which is calculated using the principles of a fixed-effects meta-analysis [30]. MR-Egger regression (which yields an intercept term to denote the presence or absence of unbalanced horizontal pleiotropy) [31] and the WME can give more robust estimates when up to 50% of the genetic variants are invalid and thus, do not meet all MR assumptions [32]. For the cognitive function outcomes results are expressed as expß-coefficients for log-transformed outcomes, which should be interpreted as % differences in the outcome for every 1-unit increase in daytime napping frequency. For the neuroimaging outcomes results are expressed as unstandardised beta coefficients to be interpreted as differences in the outcome (in cm3) for every 1-unit increase in daytime napping.

#### ii. Sensitivity analyses

a. *To ensure that our results were robust we performed all of our MR analyses additionally using a 47-SNP and 86-SNP daytime napping instrument, as described earlier. We confirmed a priori before implementing our analyses that these instruments were of adequate strength (via F-statistics)*.
b. *To mitigate potential issues with sample overlap between the discovery GWAS for daytime napping and our analytical dataset (both used UKB) we additionally performed our MR analyses using a reduced 17-SNP daytime napping instrument. This instrument consisted of the SNPs that were replicated (at P<5*10*^*-8*^) [28] *in an independent cohort (23andMe, n=541 333), as an a priori F-statistic confirmed that it was suitable for use in our MR analyses (F=67*.*1). We only performed these analyses for the cognitive function outcomes, as the overlap in samples between daytime napping and our neuroimaging analytical sample was <10% and it is possible that analyses with a 17-SNP instrument in our subsample of ∼35,000 would result in imprecise MR estimates*.

#### iii. Testing of MR assumptions

a. *Associations between genetic instrument and exposure instrumented: GWAS robust: this assumption was met, as the daytime napping variants we instrumented here have been robustly associated with this phenotype in a recent very large-scale GWAS*.
b. *No evidence of horizontal pleiotropy (no association between genetic instruments and the outcome, other than via the exposure under study): we tested this assumption by implementing MR-Egger and WME sensitivity analyses, as detailed above*.
c. *No associations between genetic variants and confounders of the relationships under study: to assess this assumption we regressed a number of common confounders on our main instrument (92 SNPs) and used a Bonferroni multiple testing correction of 0*.*05/92=0*.*0005. The list of confounders we selected was based on recent literature* [8] *and included: years of full-time education, deprivation (Townsend deprivation quintiles), smoking (ever/never/ex-smoker), physical activity (days of moderate activity for more than 10 minutes), body mass index (BMI) (kg/m*^*2*^*), alcohol consumption (1-8 times per month/16 times per month-daily/rarely or never), prevalent type-2 diabetes (No/Yes), prevalent hypertension (No=not on antihypertensive medication, Yes=on antihypertensive medication), prevalent cardiovascular disease (No/Yes)*.

## Results

### Sample characteristics

In our overall sample 53% of participants were female with a mean age of 57 years, spent an average of 15 years in full-time education and 22% were in the most deprived quintile. The mean reaction time was 555 milliseconds and the mean number of visual memory errors recorded was four, while average BMI was 27.3kg/m^2^. At baseline, there were 20 228 participants with diabetes, 29 747 with CVD, 93,092 on antihypertensive medication. Fifty per cent reported consuming alcohol between 16 times per month-daily. Participants did an average of 3.6 days of moderate physical activity for more than 10 minutes and 27% reported ever smoking. Mean hippocampal volume was 3.8cm3, while mean total brain volume was 1492cm3.

### Main MR results

#### Associations between daytime napping and total brain, and hippocampal volumes using a 92-SNP genetic instrument

As illustrated in Figure 3, IVW showed that genetic liability to daytime napping was associated with 15.80 cm3 larger total brain volume. Both MR-Egger and WME approaches indicated no unbalanced horizontal pleiotropy (Egger intercept *P*-values >0.05). The MR-Egger slope was not directionally consistent with the IVW estimate. However, the WME estimate was consistent in terms of direction and size (13.28 cm3), but did not reach conventional levels of statistical significance.

#### Associations between daytime napping and cognitive function using a 92-SNP genetic instrument

Figures 1 and 2 shows that using our main instrument we found no associations between daytime napping and reaction time or visual memory. We also found no evidence of horizontal pleiotropy using MR-Egger and WME approaches (all MR-Egger intercept *P*-values >0.05).

**Figure 1.**
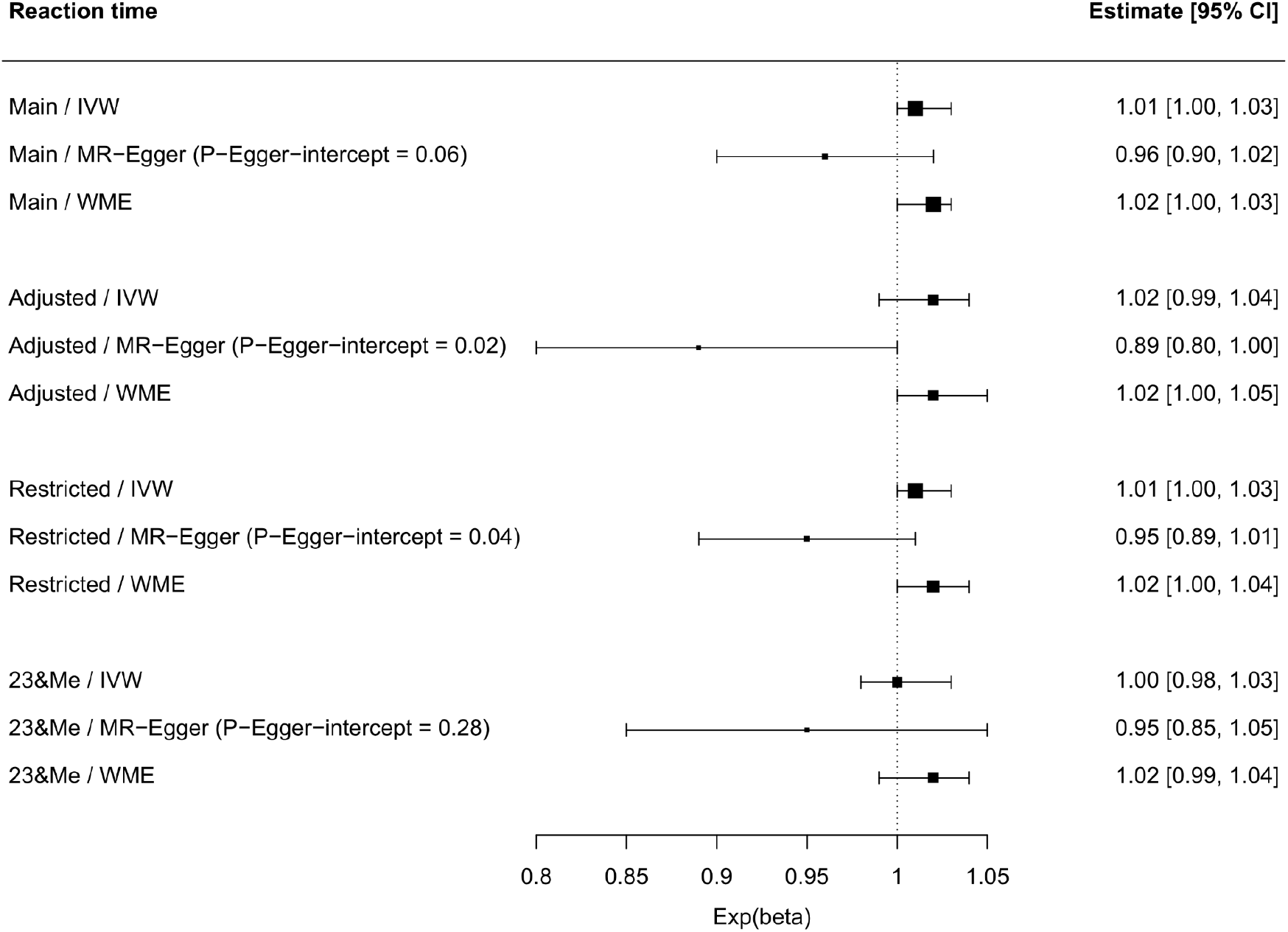
Associations between daytime napping and reaction time in UK Biobank including sensitivity analyses. *Note. n*=378 932, instrument details: Main=92-SNP main daytime napping instrument from Dashti et al, 2021, Adjusted= 47-SNP instrument adjusted for excessive daytime sleepiness, Restricted= 86-SNP instrument excluding individuals with self-reported sleep apnoea, 23&Me= 17-SNP instrument used as it has no sample overlap with UKB. IVW=inverse-variance weighted, WME=weighted median estimator, 95%CI=95% confidence interval. Exp(beta): exponentiated beta (e.g. an exponentiated beta of 1.01 in reaction time represents an estimated 1% increased/slower reaction time for every 1-unit increase in daytime napping frequency).

**Figure 2.**
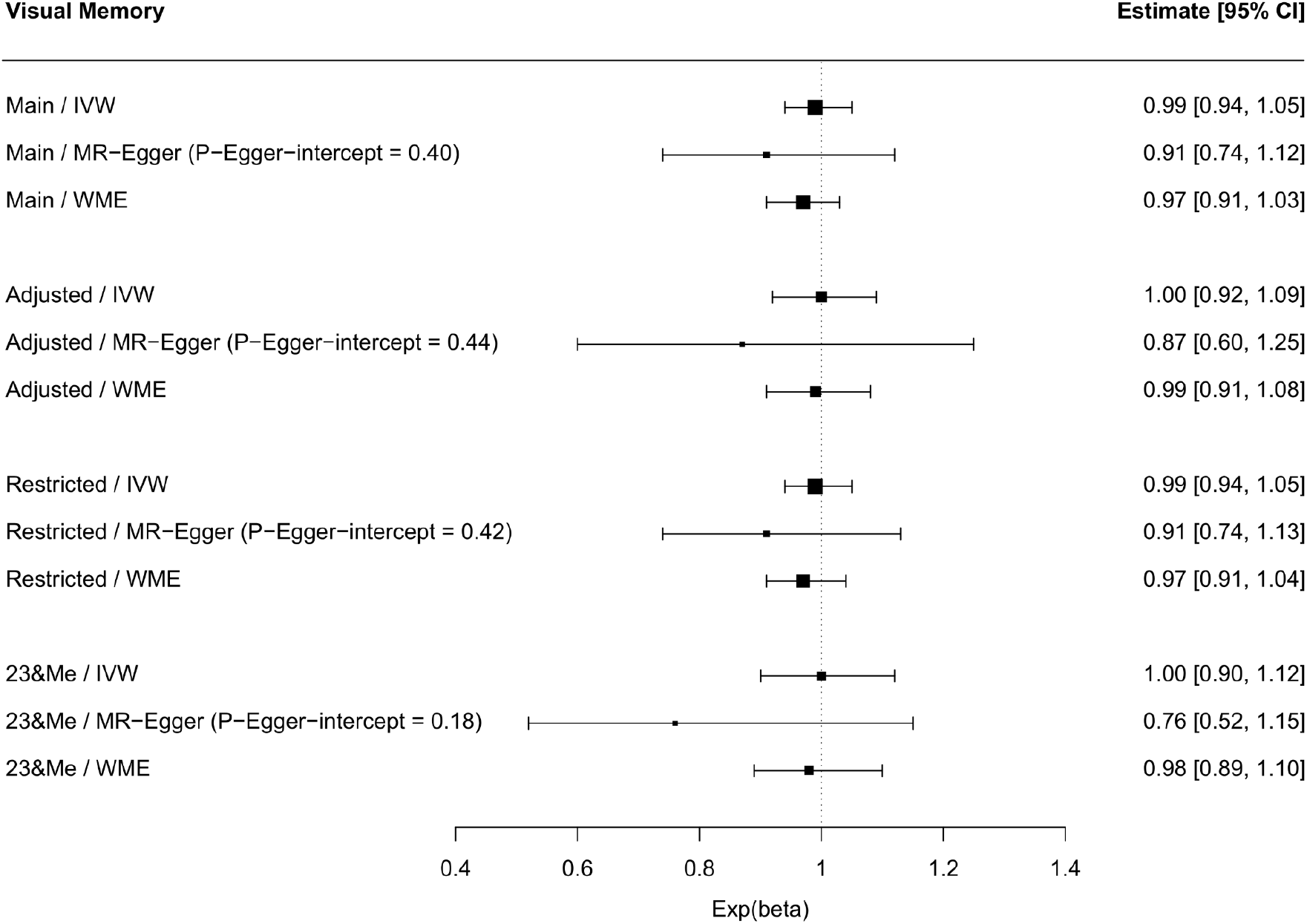
Associations between daytime napping and visual memory in UK Biobank including sensitivity analyses. *Note. n*=378 932, instrument details: Main=92-SNP main daytime napping instrument from Dashti et al, 2021, Adjusted= 47-SNP instrument adjusted for excessive daytime sleepiness, Restricted= 86-SNP instrument excluding individuals with self-reported sleep apnoea, 23&Me= 17-SNP instrument used as it has no sample overlap with UKB. IVW=inverse-variance weighted, WME=weighted median estimator, 95%CI=95% confidence interval. Exp(beta): exponentiated beta.

**Figure 3.**
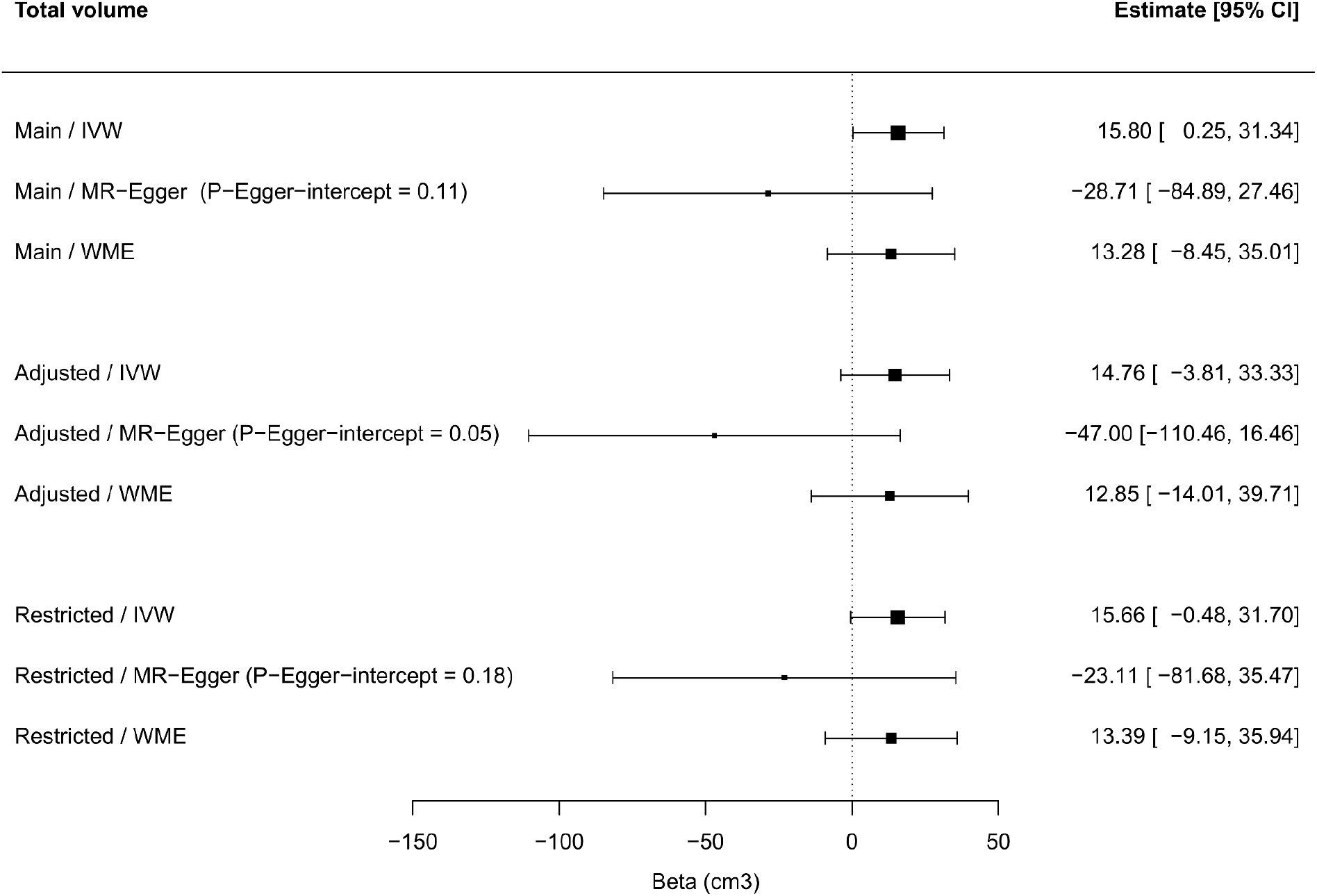
Associations between daytime napping and total brain volume in UK Biobank including sensitivity analyses. *Note. n*=35,080, instrument details: Main=92-SNP main daytime napping instrument from Dashti et al, 2021, Adjusted= 47-SNP instrument adjusted for excessive daytime sleepiness, Restricted= 86-SNP instrument excluding individuals with self-reported sleep apnoea, 23&Me= 17-SNP instrument used as it has no sample overlap with UKB. IVW=inverse-variance weighted, WME=weighted median estimator, 95%CI=95% confidence interval.

### Sensitivity analyses

#### Associations between daytime napping and total brain, and hippocampal volumes using a 47- and 86-SNP genetic instrument

When we used a 47-SNP daytime napping instrument (adjusted for excessive daytime sleepiness) the associations with total brain volume were consistent in terms of size and direction with our main results (Figure 3). This was very similar for associations between the 86-SNP daytime napping and total brain volume (Figure 3). However, potentially due to lower total power (particularly in terms of the variance explained (R2) in daytime napping by these reduced instruments) these estimates had wider 95% CIs around them. In line with our main results above, we observed no association between a 47-SNP daytime napping instrument (excluding individuals with self-reported sleep apnoea) and hippocampal volume, or an 86-SNP instrument and hippocampal volume (Figure 4). MR-Egger detected the presence of unbalanced horizontal pleiotropy using the 47-SNP instrument. Therefore, we excluded the SNP that was most strongly associated with total brain volume (rs301817), reran our MR analyses and the MR-Egger intercept *P*-value was >0.05. The IVW and WME estimates, as well as the MR-Egger slope remained very similar (and all estimates still crossed the null) and we have not presented them here. There were no other issues with unbalanced horizontal pleiotropy, as per the MR-Egger and WME results.

**Figure 4.**
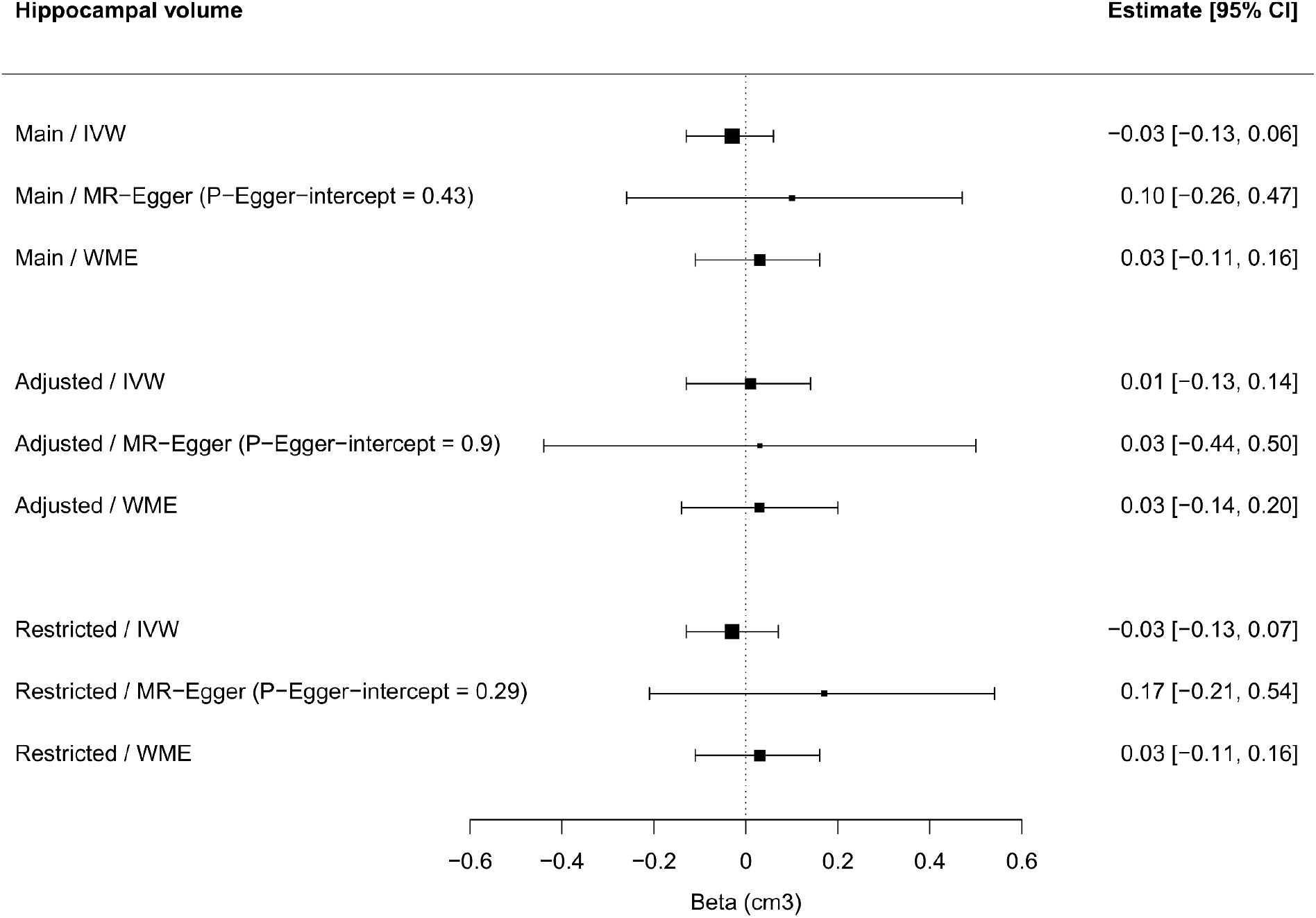
Associations between daytime napping and hippocampal volume in UK Biobank including sensitivity analyses. *Note. n*=35,080, instrument details: Main=92-SNP main daytime napping instrument from Dashti et al, 2021, Adjusted= 47-SNP instrument adjusted for excessive daytime sleepiness, Restricted= 86-SNP instrument excluding individuals with self-reported sleep apnoea, 23&Me= 17-SNP instrument used as it has no sample overlap with UKB. IVW=inverse-variance weighted, WME=weighted median estimator, 95%CI=95% confidence interval.

#### Associations between daytime napping and cognitive function using a 47- and 86-SNP genetic instrument

As results presented in Figure 1 and 2 suggest, sensitivity analyses using the 47-SNP instrument also showed no associations with reaction time or visual memory. Similar results emerged for the 86-SNP instrument with no evidence of associations with either of the two cognitive function measures. For reaction time the MR-Egger intercept *P*-value indicated the presence of unbalanced horizontal pleiotropy using both the 47- and 86-SNP instruments. Thus, we excluded one SNP that was the most strongly associated with reaction time (rs2099810), re-ran our MR analyses and the MR-Egger intercept had *P*>0.05. The MR-Egger slopes, as well as the IVW and WME results, remained unchanged and are therefore not presented. However, we did not detect any issues with horizontal pleiotropy for visual memory, with both MR-Egger intercept *P*-values >0.05.

#### Association between daytime napping and cognitive function using a 17-SNP instrument with no sample overlap

Using this restricted instrument to ensure no overlap between our exposure and outcome samples, across all three MR approaches we observed no associations with reaction time or visual memory. MR-Egger detected no issues with unbalanced horizontal pleiotropy (*P*>0.05). Results are presented in Figure 1 and 2.

### Testing MR Assumption III

#### Associations between our main 92-SNP daytime napping genetic instrument and common confounders

After a Bonferroni correction we observed that 12 variants were associated with education, two with deprivation, four with smoking, two with physical activity, 19 with BMI, one with alcohol consumption, three with diabetes, eight with hypertension and one with CVD. We present these associations in **Supplementary Table 3**.

## Discussion

Using a comprehensive Mendelian randomisation design, we found an association between genetic liability to self-reported habitual daytime napping and larger total brain volume but not hippocampal volume, reaction time, or visual memory in the UK Biobank. To our knowledge, no prior studies have used MR to try to disentangle the relationship between daytime napping and cognitive and structural brain outcomes.

The association found between habitual daytime napping and larger total brain could suggest that this behaviour provides some protection against neurodegeneration. Measures of brain volume have been used as proxies of neurodegeneration [33] and are thought of as strong candidates to explain some of the variations related to cognitive ageing [13, 14]. Reductions in brain volume are expected throughout the lifespan [34], but this process is accelerated in people with cognitive decline and neurodegenerative diseases [35]. Crucially, it is proposed that sleep deficits could be related to these structural changes [36]. For example, several neuroimaging studies have found lower brain volume in people with sleep problems, such as insomnia [37, 38] and poor sleep quality [39]. Moreover, it has been suggested that sleep disturbances may be risk factors for neurodegenerative disorders [33, 40] by promoting processes such as inflammation and synaptic damage [41]. In line with this, recent MR studies found that daytime sleepiness was associated with higher Amyotrophic Lateral Sclerosis (ALS) risk [20, 21] and sleep efficiency was associated with lower AD risk [20]. In a similar vein, Anderson et al. (2021) found suggestive evidence that reduced daytime napping is associated with higher AD risk. These studies suggest that inadequate sleep could lead to neurodegeneration and that daytime napping could compensate for poor nocturnal sleep.

Our finding of larger total brain volume in relation to habitual daytime napping was found only using the IVW estimate with our main genetic instrument (92 SNPs). However, we wish to emphasise that the IVW estimate in the adjusted (47 SNPs; 14.76cm^3^) and the restricted (86 SNPs; 15.66cm^3^) instruments were almost identical to the estimate using our main instrument (15.80cm^3^). These additional instruments were also consistent in terms of direction. Moreover, we predict that more precise estimates, with narrower confidence intervals, may be observed if we were to replicate these analyses with the entire MRI sample when it becomes available (≈100,000).

We also expected to find that habitual daytime napping would be associated with hippocampal volume. Our hypothesis was based on the fact that the hippocampus, as a brain structure that plays a crucial role in memory [42], could be a useful proxy of the variations in memory performance reported to be associated with daytime napping [43–45]. However, we did not find this association, nor an association between genetic liability to habitual daytime napping and visual memory performance. Previous studies have reported mixed findings for sleep phenotypes and hippocampal volume, with a number of studies revealing that people with sleep problems have reduced hippocampal volume [46–50], while other studies report no associations [51–53]. In line with our results, a recent cross-sectional analysis in the UKB revealed that napping was not related to hippocampal volume [54].

We were surprised by the lack of a causal link between daytime napping and reaction time, or visual memory, given the evidence of cross-sectional associations between daytime napping and cognitive outcomes [7, 8], and the relationship between cognitive function and AD [55]. However, we found no evidence to support this hypothesis. More reliable cognitive measures may be required to identify these effects. In this regard, our results may be influenced by test characteristics (e.g., task sensitivity and difficulty, timing, or instructions) [3]. Furthermore, UKB cognitive assessments are not standardised and were designed specifically for this cohort. Nonetheless, it is worth mentioning that we examined the association between genetic liability to habitual daytime napping and cognitive function and not the effect of taking a nap before performing a cognitive test. In addition, it is important to establish that, despite these limitations, UKB cognitive data is a valuable resource for researchers seeking determinants of cognitive function [27].

Moreover, individual differences in the experiences with napping, for example, the presence of sleep apnoea [56] and daytime sleepiness [3], may affect the degree of the cognitive benefit generated by naps. In this regard, we partitioned the daytime napping instrument into two sub-instruments (one excluding individuals who had sleep apnoea and the other adjusting for excessive daytime sleepiness). Still, no evidence of associations between self-reported daytime napping and reaction time, or visual memory was found. However, other factors such as slow waves production, the quality of the prior sleep period or the presence of sleep inertia could also influence napping restoration [3], which could lead to different effects on cognition. The association between napping and cognitive function may also be influenced by depression, as the frequency of napping has been associated with depressive symptoms [7, 57, 58]. Also, the relationship between depression and cognition is well established [59, 60].

In addition, we only analysed the frequency of napping. However, observational studies have shown that the length and timing of naps could also affect cognitive function [11]. Unfortunately, information on these dimensions is not available in UKB. Regarding length, previous studies reported that, unlike long naps, the beneficial effects of brief naps are evident almost immediately after waking but last for a shorter period of time [11]. Nap’s timing also determines its effect on cognition, with the post-lunch dip period being the most favourable time to take a nap to overcome the temporary drop in alertness and performance evidence during this period [61].

To validate our MR findings, it was checked that the three core assumptions that underlie MR were met. Assumption I was met as we instrumented the best available genetic variants as they have been robustly associated with daytime napping in a recent large-scale GWAS [28]. MR-Egger and WME sensitivity analyses were implemented to check assumption II. No evidence of horizontal pleiotropy was found, which corroborates that the association between our genetic variants (for the exposure) and outcomes were only via the exposure under study. Finally, assumption III was tested by performing regressions between our genetic instruments and unobserved confounders, and we found that some of the variants were associated with common confounders. These associations should be further investigated, as they may constitute vertical, rather than horizontal pleiotropy.

### Limitations

Limitations of the study should be noted. First, our exposure and cognitive outcome samples overlapped by 77%. However, sensitivity analyses using a reduced 17-SNP daytime napping instrument, replicated by the GWAS authors [28] in an independent cohort (23andMe), confirmed that it was suitable for use in our MR analyses. Using this reduced instrument, we observed no associations with reaction time or visual memory. Second, participants were only white European; future work should examine if these findings are replicated in other ancestries. Third, future instruments for the length and timing of daytime napping are necessary. Fourth, another limitation of our study was the self-report nature of the exposure under study, but napping is notoriously difficult to measure using objective methods. However, in UKB there was consistency between self-reported sleep measures and accelerometer-derived daytime inactivity duration, which increases confidence in the SNPs for daytime napping.

## Conclusions

In summary, our Mendelian randomisation study of daytime napping and cognitive/structural brain outcomes suggests an association between genetically-instrumented daytime napping and larger total brain volume, but not hippocampal volume, reaction time, or visual memory. This study improves our knowledge of the impact of habitual daytime napping on brain health which is essential to understanding cognitive impairment in the ageing population. The lack of evidence for an association between napping and hippocampal volume and cognitive outcomes in the present study may indicate that other brain areas and cognitive outcomes (e.g. alertness) may be affected by habitual daytime napping and should be studied in the future. These findings further our understanding of the relationship between daytime napping frequency and cognitive function and structural brain outcomes and elucidate the importance of using different measures to better understand how sleep relates to brain health.

## Data Availability

The UK Biobank data are publicly available to all bona fide researchers at
https://www.ukbiobank.ac.uk

## Declarations

### Ethics approval

Ethics approval is not needed as this work was conducted under the approved UK Biobank project number 71702.

### Author contributions

Literature search: VP. Study design: VG. Data analysis: VG. Data interpretation: VG, VP, HSD. Writing the manuscript: VG and VP. Revision of the manuscript: VG, VP, HSD. Final approval of the manuscript: VG, VP, HSD.

### Data Availability

The UK Biobank data are publicly available to all bona fide researchers at https://www.ukbiobank.ac.uk.

## Acknowledgements

This work was conducted under the approved UK Biobank project number 71702. We thank all UKB researchers and volunteers.

