## Supplementary Figure S1 for "Is there an association between daytime napping, cognitive function and brain volume? A Mendelian randomisation study in the UK Biobank"

Figure S1. SNPs selection process

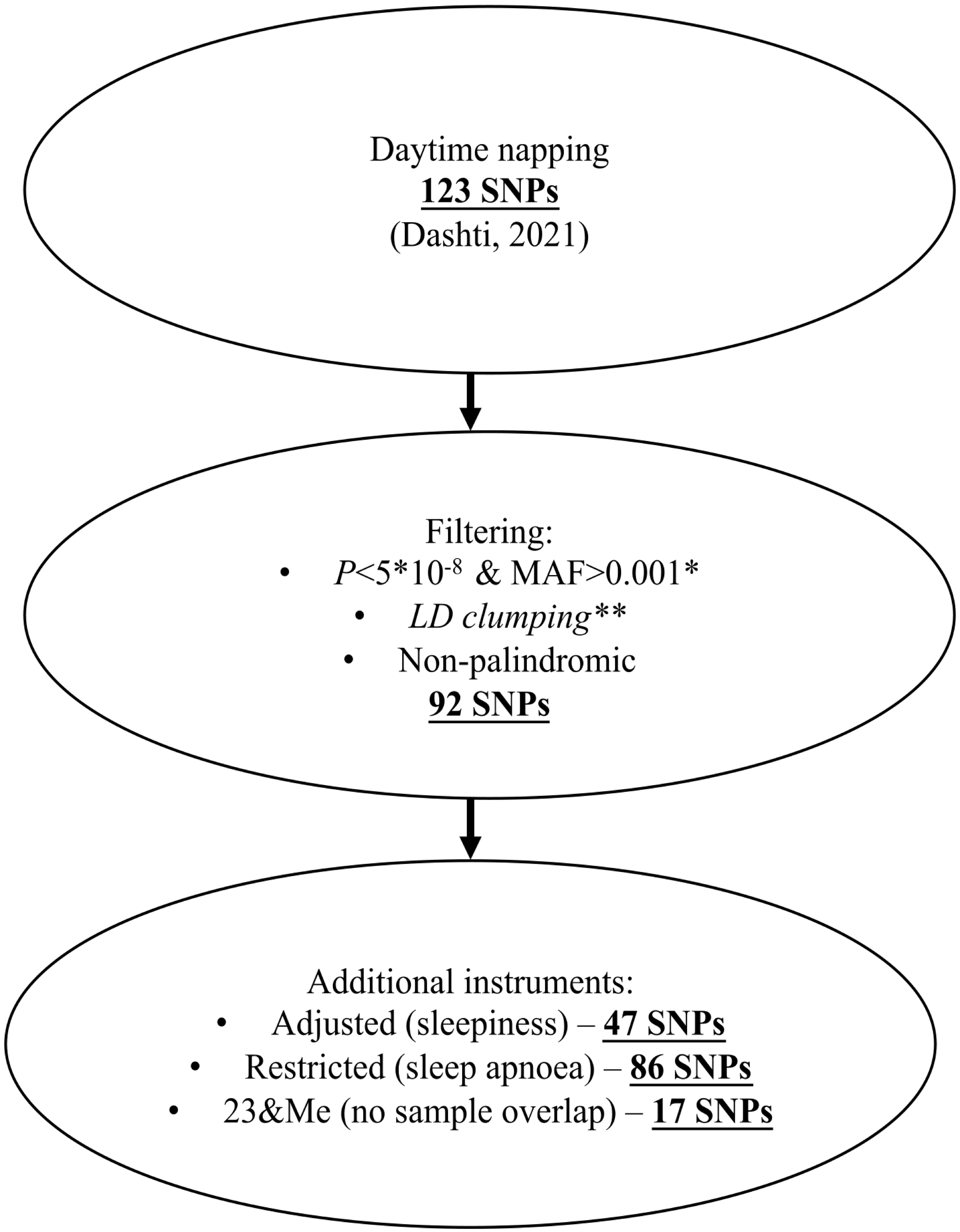


*Note. *MAF filtering at 0.001 done by GWAS authors; **passed genetic QC and confirmed to be independent at LD clumping thresholds of $r^2<0.\ 01$, within a 250 kb window, using 1000 Genomes CEU data.*
